# Predicting Mental and Neurological Illnesses Based on Cerebellar Normative Features

**DOI:** 10.1101/2024.12.10.24318590

**Authors:** Milin Kim, Nitin Sharma, Esten H. Leonardsen, Saige Rutherford, Geir Selbæk, Karin Persson, Nils Eiel Steen, Olav B. Smeland, Torill Ueland, Geneviève Richard, Aikaterina Manoli, Sofie L. Valk, Dag Alnæs, Christian F. Beckman, Andre F. Marquand, Ole A. Andreassen, Lars T. Westlye, Thomas Wolfers, Torgeir Moberget

## Abstract

Mental and neurological conditions have been linked to structural brain variations. However, aside from dementia, the value of brain structural characteristics derived from brain scans for prediction is relatively low. One reason for this limitation is the clinical and biological heterogeneity inherent to such conditions. Recent studies have implicated aberrations in the cerebellum – a relatively understudied brain region – in these clinical conditions. Here, we used machine learning to test the value of individual deviations from normative cerebellar development across the lifespan (based on trained data from >27k participants) for prediction of autism spectrum disorder (ASD) (n=317), bipolar disorder (BD) (n=238), schizophrenia (SZ) (n=195), mild cognitive impairment (MCI) (n=122), and Alzheimer’s disease (AD) (n=116). We applied several atlases and derived median, variance, and percentages of extreme deviations within each region of interest. Our results show that lobular and voxel-wise cerebellar data can be used to discriminate healthy controls from ASD and SZ with moderate accuracy (the area under the receiver operating characteristic curves ranged from 0.56 to 0.64), The strongest contributions to these predictive models were from posterior regions of the cerebellum, which are more strongly linked to higher cognitive functions than to motor control.

## Introduction

Clinical heterogeneity and complex pathobiological mechanisms impede the discovery of reliable biomarkers for many neurological and – especially – psychiatric disorders, thereby complicating precise clinical decision-making and treatments. Over the last two decades, there has been a trend in the development of neuroimaging-based tools and machine learning for prognosis and diagnosis of psychiatric disorders (1,2) and neurological illnesses (3). Neuroimaging-based prediction studies on autism spectrum disorder (ASD), bipolar disorder (BD), and schizophrenia (SZ) have reported a wide range of accuracies, underscoring the limitations associated with small samples, including poor generalization performance (4,5). Of note, prediction studies on dementias show greater promise for clinical usage in both Alzheimer’s disease (AD) (3) and mild cognitive impairment (MCI).

Notably, the majority of these prediction studies (4–6) have focused on cerebral features, perhaps reflecting a “cortico-centric bias” in the literature (7). Nonetheless, disruptions in the cerebellum have been hypothesized to contribute to various clinical conditions, such as childhood psychiatric symptoms (8), AD (9), SZ (10), and ASD (11–13). Indeed, patient studies have shown that abnormalities in cerebellum can exert a significant influence on motor, cognitive, and emotional functions (14–16), yet, there is little exploration on the role of the cerebellum in predicting and classifying mental and neurological illnesses. Using a normative modelling approach, we recently demonstrated significant deviations from normal cerebellar developmental across the lifespan in ASD, MCI, AD, BD, and SZ (17). While these individual-level deviations revealed substantial cerebellar heterogeneity among individuals with the same disorder, the value of these cerebellar features with respect to classifying these disorders remain uncertain.

In this study, we addressed this gap by performing a set of predictions of ASD, MCI, AD, BD, and SZ, using MRI-based cerebellar features and cross-validated machine learning classifiers. We applied lobular and voxel-wise normative models (17) and aggregated the median, variance and percentage of extreme deviations across atlases (18,19). Finally, for models that were able to meaningfully differentiate between patients and health controls, we identified cerebellar regions that contributed the most to the prediction.

## Methods

### Sample

The study consisted of healthy controls from the test set the cerebellar lifespan normative model (17) (n=26.985, 53% females), and the clinical samples (n=1.757, 30% females) (Figure 1A and Supplementary Table 2). Individuals without diagnoses were matched to the clinical datasets of patients with AD, ASD, BD, MCI, and SZ (Table 1) using nearest neighbor matching based on exact matches of sex and scanning site with age as implemented in *MatchIt* (20). The clinical datasets were obtained from the ABIDE, ADNI, AIBL, DEMGEN, and TOP cohorts. Information about each cohort and studies can be found in the corresponding publications (Supplementary Table 1). If participants were scanned at several timepoints, only baseline scans were chosen for this study. Individuals who withdrew from the studies or lacked essential demographic information and T1-weighted MRI data were excluded from the analyses.

**Figure 1.**
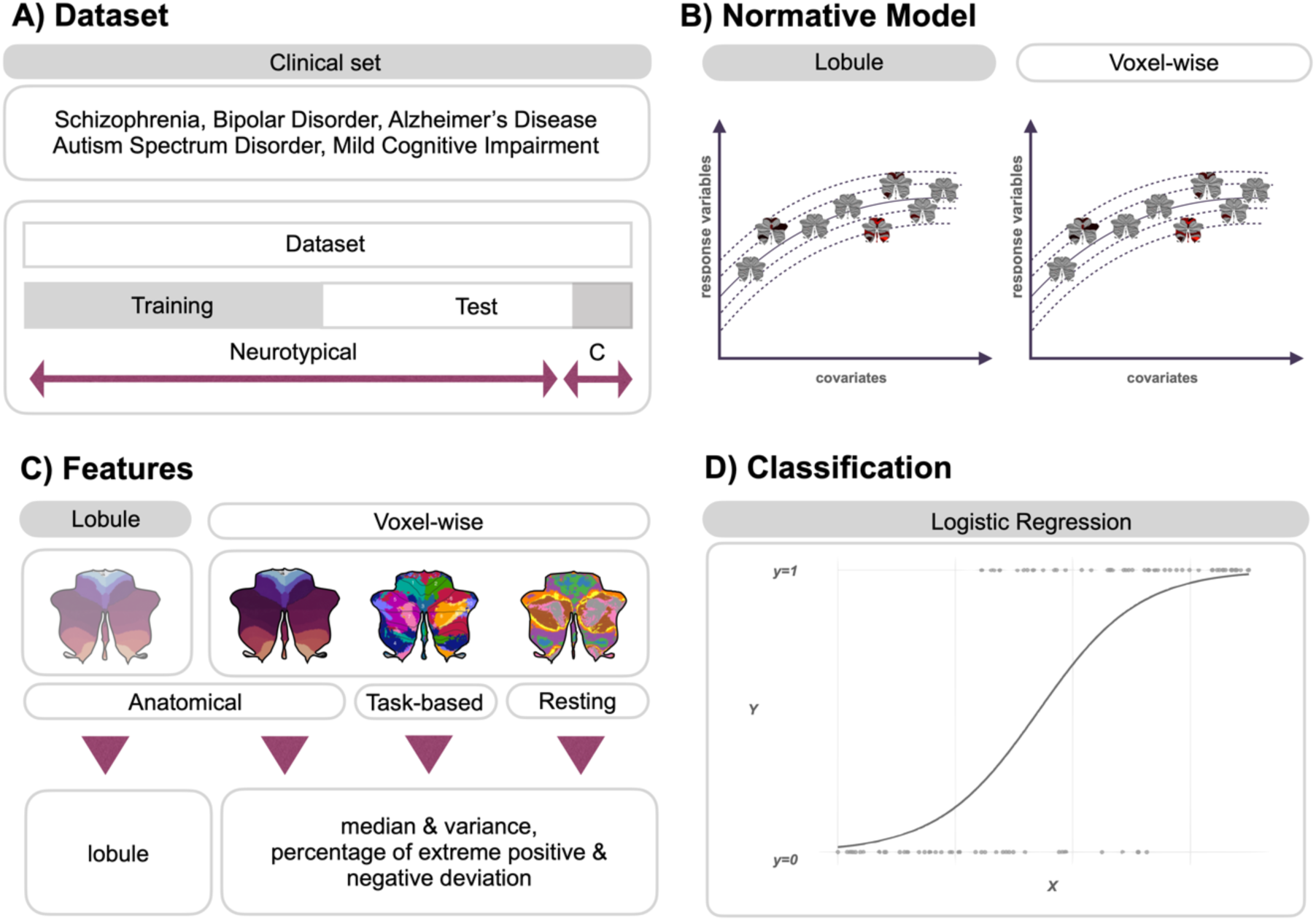
Overview of Predicting Mental and Neurological Illnesses. (A) The study incorporated five clinical datasets. (B) Individuals without a diagnosis were divided into training and testing sets to evaluate the cerebellar normative models, which were prepared in both lobular and voxel-wise features. (C) The analysis utilized six distinct types of features, median, variance, and percentage of extreme positive and negative deviation, alongside lobular volume. (D) Logistic regression algorithm was employed to determine the likelihood of a clinical diagnosis in an individual.

**Table 1.** Matched sample description and demographics.

|  |  | N (Participants) | N (Scanners) | Age (Mean, S.D.) | Sex (%F: %M) |
| --- | --- | --- | --- | --- | --- |
| Matched<br>HC | Alzheimer's Disease | 116 | 13 | 71.72(7.12) | 55:45 |
|  | ASD | 317 | 25 | 15.96(7.45) | 17:83 |
|  | Bipolar Disorder | 238 | 3 | 33.06(10.50) | 55:45 |
|  | Mild Cognitive<br>Impairment | 122 | 3 | 65.62(9.91) | 42:58 |
|  | Schizophrenia | 195 | 3 | 30.13 (8.15) | 41:59 |
| Clinical | Alzheimer's Disease | 116 | 13 | 73.11(7.60) | 55:45 |
|  | ASD | 317 | 25 | 12.35(4.42) | 17:83 |
|  | Bipolar Disorder | 238 | 3 | 31.61(11.40) | 55:45 |
|  | Mild Cognitive<br>Impairment | 122 | 3 | 67.25(9.27) | 42:58 |
|  | Schizophrenia | 195 | 3 | 28.29 (9.45) | 41:59 |

### Lobular-level processing

The T1-weighted images were skull-stripped using the FreeSurfer 5.3 auto-recon pipeline (21) and reoriented to the standard FSL orientation using the *fslreorient2std* (22). Linear registration was performed using *flirt* (23), which employed linear interpolation (with six degrees of freedom) and the default 1 mm FSL template (version 6.0). The borders were cropped at coordinates [6:173, 2:214, 0:160] to minimize their size without removing brain tissue. Finally, the voxel intensity values of all brain images were normalized to the range of [0,1], adjusting the intensity values of each voxel to a standardized scale.

To segment the cerebellum, we utilized the ACAPULCO algorithm (24), part of ENIGMA Cerebellum Volumetric Pipeline, which is a cerebellum parcellation algorithm based on convolutional neural networks. This algorithm delivers fast and precise quantitative in-vivo regional assessment of the cerebellum. As part of the algorithm, the images were corrected for inhomogeneity by N4 correction method (25) and registered to the 1mm isotropic ICBM 2009c template in MNI space using the ANTs registration suite (26). The ACAPULCO algorithm is based on 15 expert manual delineations of an adult cohort (27). It achieves per-voxel labelling and employs post-processing of the parcellation to correct for mislabeling and for accurate segmentation. ACAPULCO segments the cerebellum into 28 cerebellar lobules and computes the volume (mm^3^) for each region. These regions include bilateral Lobules I–VI; Crus I and II; Lobules VIIB, VIIIA, VIIIB, and IX-X; Vermis VI, VII, VIII, IX, and X; and Corpus Medullare (CM). To ensure data quality, participants with extreme outliers (2.698 s.d. above or below the mean) (28) in more than two lobules based on automated quality control measures, were excluded. We set the threshold at two lobules because the differences between one and two lobules was not significant (see Supplementary Methods for detailed information for quality control).

### Voxel-level processing

We used SUIT version 3.4 (Spatially Unbiased Infratentorial Toolbox) (29) to segment cerebellar grey and white matter voxel-based morphometry (VBM) maps. SUIT leverages the outputs from ACAPULCO, an MNI-aligned T1 image (29,30), and an average mask derived from a randomly selected group of 300 individuals without a diagnosis. After segmentation, the grey matter maps were normalized for standardized comparison and re-sliced to align them with the MNI152 template. Additionally, the grey matter maps were modulated by the Jacobian to preserve the value of each voxel in proportion to its original volume. This Jacobian modulation ensured that the values of the original volume were proportionally maintained.

### Normative modelling

We used a publicly available cerebellar normative model, estimated using >27 participants (17) which is implemented in the PCNtoolkit package (version 0.24) (31,32). This normative model encompasses cerebellar volumes and voxel-wise intensity while including sex, age, and scanning-site as covariates (Figure 1B).

To analyze the data, we employed Bayesian Linear Regression (BLR) with the likelihood warping method (33), incorporating the ‘sinarcsinsh’ transformation (34,35), to handle non-linear basis functions and non-Gaussian predictive distributions for large datasets (34). Scanning-site was accounted for as a fixed effect (36,37). The normative model provides point estimates and evaluation metrics such as explained variance, mean squared log-loss, skew, and kurtosis (35). These evaluation metrics were calculated in the test set, which did not include clinical cohorts. Extreme deviations were defined as |z| > 1.96, corresponding to the most extreme 5% of cases in both directions in the reference cohort.

### Feature engineering

Voxel-wise normative models were utilized to map deviation profiles onto existing atlases (see Supplementary Methods). Three existing atlases were selected: 28 cerebellar anatomical regions, 10 regions of interest (ROI) from the multi-domain task battery (MDTB) (19), and 17 ROI from resting-state connectivity (18,38). For each region of interest delineated by these atlases, we computed three key statistics: the median, variance and percentage of extreme deviations (Figure 1C). To quantify the extremes in deviation, we also calculated the proportion of voxel-wise deviations that exceeded the established threshold of |z| > 1.96, denoting both extreme positive and negative deviations. This proportion was determined by dividing the count of such extreme deviations by the total voxel count within the corresponding region of interest. Variance has previously been used to examine the structural heterogeneity among patients in SZ (39,40). Unlike percentage of extreme deviation (|*Z*| > 1.96) that has been used in past normative studies (41–43), variance assesses the dispersion within the region, capturing the regionally heterogeneous spread within patients.

### Model training and evaluation

Machine learning models employing logistic regression (LR) were used to build prediction models (Figure 1D). In addition, results from the random forest (RF) algorithm from the scikit-learn library version 1.2.2 (44) and the eXtreme Gradient Boosting (XGBoost) library version 1.7.3 (45) can be found in the Supplementary Figure 3. RF is a non-parametric supervised learning method that addresses over-fitting by combining decision trees into a single outcome, effectively balancing the bias-variance trade-off. XGBoost is an open source library to implement advanced gradient boosting algorithms (45).

The features engineered from three atlases separately ran as inputs for the algorithm. We developed various machine learning models using deviations from the normative models and utilized their median, variance and percentage of extreme deviation onto the existing atlases as features. To evaluate the model’s performance in held-out test data, we conducted a stratified five-fold cross-validation and used the area under the receiver operating characteristic curve (AUROC) as the primary performance metric. Additionally, we calculated precision, recall, sensitivity, specificity, balanced accuracy, and the area under the precision-recall curve.

### Permutation testing

We used permutation testing to assess whether the AUROCs achieved by our model was different from chance level performance. To achieve this, we shuffled the diagnosis labels randomly 1000 times, for each permutation calculating an AUROC. For significance testing, the original AUROC was compared to the distribution of permuted AUROC values. If the original AUROC falls within the extreme ends of the permutation distribution (p < 0.05), it is considered statistically significant. We applied an identical approach for the lobular volume features. The comparison between models utilized an approach similar to that outlined in Supplementary Figure 2-3, wherein the previously calculated shuffled AUROC values were used. We calculated the difference in true AUROC scores, as well as the AUROC differences from 1000 permuted datasets, between the two models. Subsequently, we compared the true score and the permuted scores to assess statistical significance.

### Feature importance ranking

We assessed feature importance based on logistic regression coefficients to highlight their influence on the predictions. The coefficients from the model directly infer the relative importance of each feature, thus facilitating interpretation. The magnitude of the coefficient indicates the strength of the effect a feature has on the prediction, while the sign (positive or negative) indicates the direction of the effect. Figure 3 illustrates the summary plot of the standardized feature importance, emphasizing the key features that have the greatest influence (see Supplementary Figure 1 for all feature importance).

## Results

We conducted a comprehensive analysis at the lobular and voxel-wise level employing a variety of models (Figure 1C). The voxel-wise model calculations included variance, median, and percentage of deviations across 143k voxels, organized into 28 ROIs for the anatomical atlas, 10 ROIs for the task-based atlas, and 17 ROIs for the resting-state atlas.

Permutation testing revealed significant predictions for ASD and SZ (AUROC values ranging from 0.56 to 0.64), using various models based on deviations from the cerebellar normative model (Figure 2). Prediction performance for MCI, AD and BD were not above chance levels. For SZ, the most predictive models were those centered around median and variance measures summarized within ROIs for the voxel-wise models. In contrast, for ASD, models based on the lobular volumes and voxel-wise variance within ROIs were found to be the most predictive. No notable differences between models based on different parcellations were found (Supplementary Figure 2). Furthermore, AUROC scores from RF predictions showed similar patterns of the logistic regression, underscoring the consistency of these techniques (Supplementary Figure 3).

**Figure 2.**
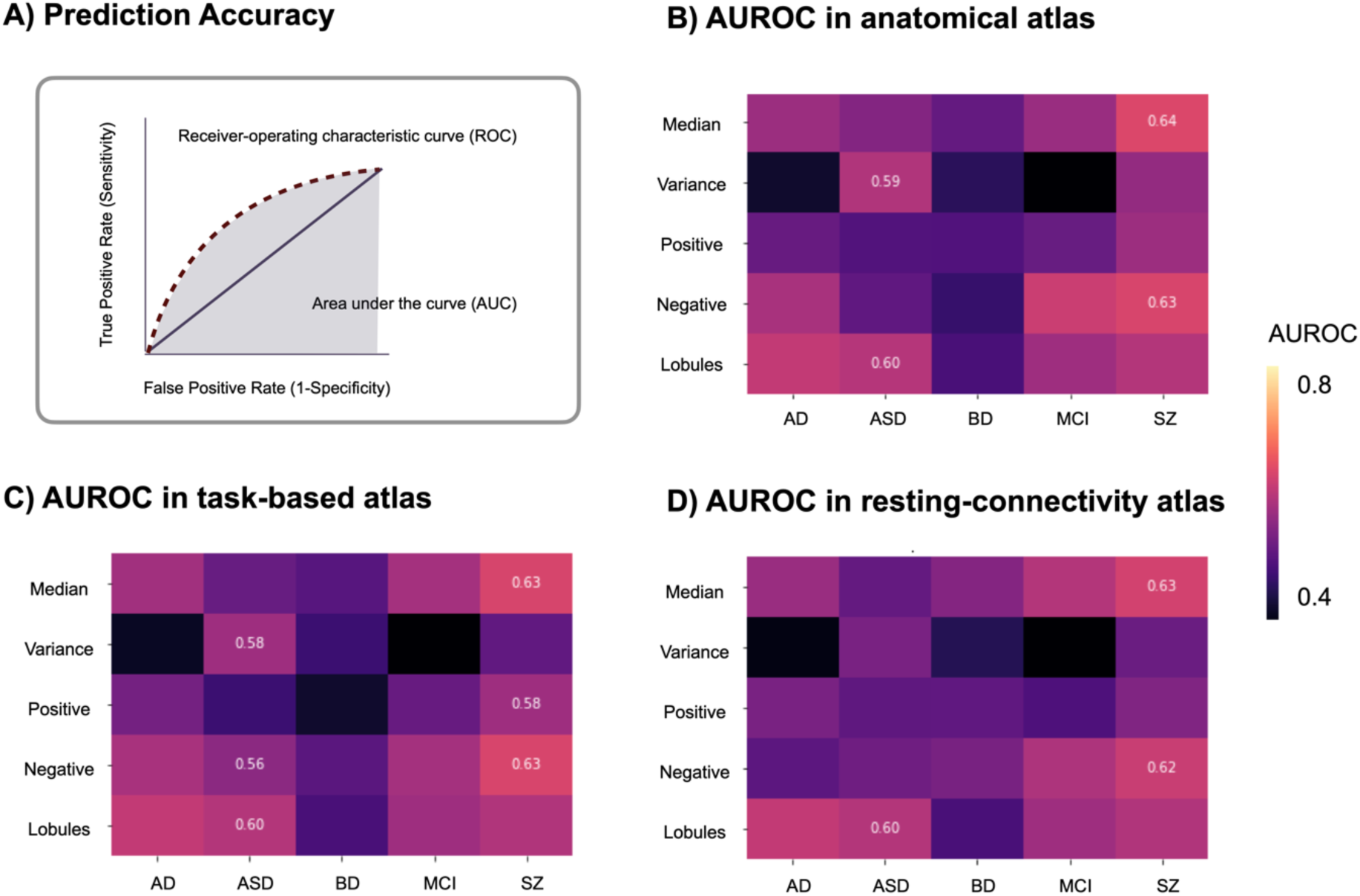
Cerebellar features moderately predict ASD and SZ. (A) Information from the anatomical atlas (28 regions), task-based (10 regions) or resting-state (17 regions) are compiled into features that were used as predictors by the logistic regression model to make predictions. The area under the receiver operating characters curve (AUROC) serves as an important measure in evaluating the performance of a binary classifier, representing a trade-off between the classifier’s sensitivity (true positive rate) and specificity (true negative rate). The reliability and robustness of the AUROC were assessed by computing it over 1,000 permutations, which aids in determining whether the classifier’s performance is statistically significant or due to random chance. (B-D) The values that survived multiple comparison are shown.

Figure 3 presents the feature importance weights in a logistic regression model used to analyze SZ and ASD. In SZ, significant negative deviation percentages were found in the vermis IX and Left IV regions. In task-based functional areas, regions associated with verbal fluency, word comprehension and mental arithmetic (region 9) and autobiographical recall, visual letter recognition, interference resolution (region 10) were notable. From the resting-state atlas, limbic A (region 10) and somatomotor A (region 3) emerged as important. For median in SZ, the anatomical regions Right I-III and vermis VIII were highlighted. Using task-based atlases, the top predictive regions were functionality linked to divided attention (region 5) and right-hand movement (region 2). Predictive models using an atlas based on resting-state atlas highlighted Visual B (region 2) and limbic A (region 10).

**Figure 3.**
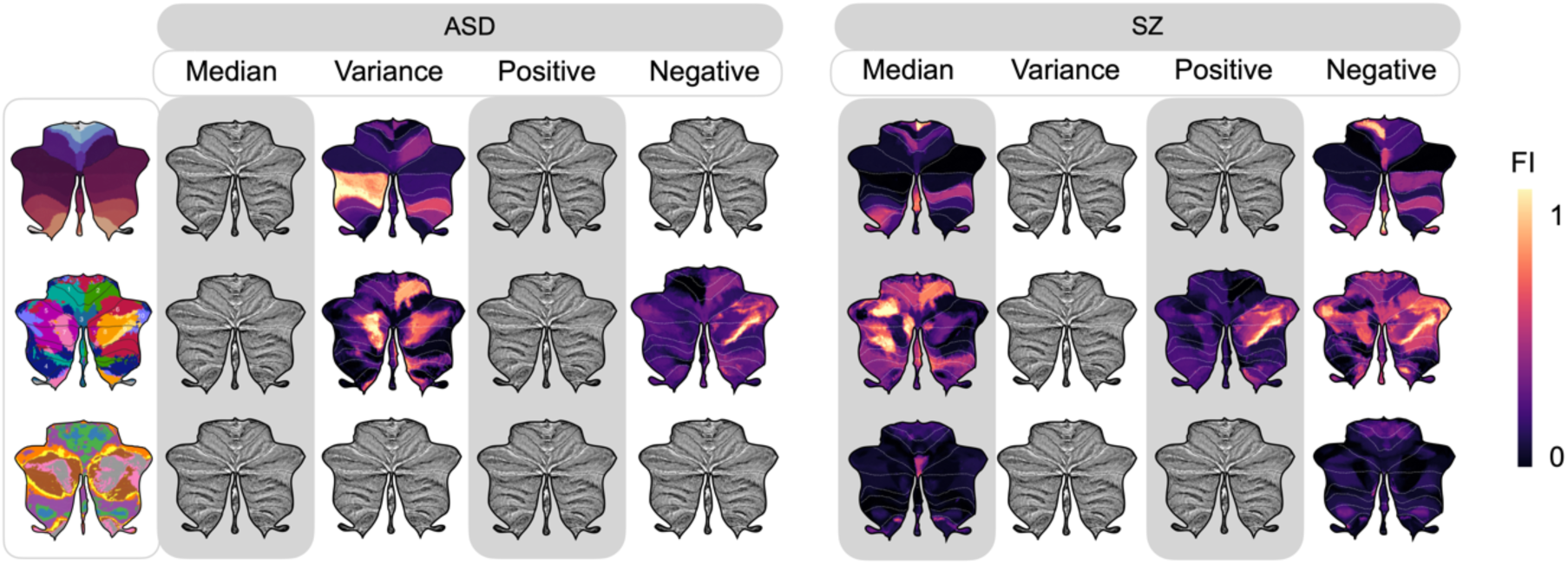
Different regions show distinct importance across atlases in ASD and SZ. The feature importance (FI) values derived from logistic regression reveal the contribution of each specific cerebellar region to predictive modelling, relative to average prediction outcomes. FI values accentuate distinct cerebellar regions with unique predictive capabilities as identified in lobules, anatomical, task-based, and resting-state atlases through voxel-wise analysis. Features that remained significant after adjustments for multiple comparisons of AUROC are shown.

In ASD, predictive models based on variance (summarized within ROIs) revealed the most significant contributions from posterior cerebellar regions of Left VIIB and Left Crus II while models based on lobular volumes features point to Right VI and Left Crus II. Using the task-based functional atlas, the most predictive regions were functionally linked to narrative, emotion, language processing (region 7) and right-hand movement, motor planning, divided attention (region 2). Complementary insights and detailed rankings of feature importance are available in Supplementary Tables 3-7.

## Discussion

This study aimed to test the predictive power of deviations from normal cerebellar anatomy with respect to classifying mental and neurological disorders and yielded two main findings. First, we demonstrated that cerebellar features offer moderate power for prediction for ASD and SZ but did not reliably distinguish healthy controls from patients with BD, MCI or AD. Second, feature importance analyses showed that posterior regions of the cerebellum, known for its contributions in cognitive processes (15) were the most important predictors of ASD and SZ.

Our study reveals that features derived from lobular and voxel-wise normative model possess moderate predictive capabilities in ASD and SZ. This is in line with our previous study which reported small to medium case-control differences in normative cerebellar anatomy for both ASD and SZ (17). However, it is worth mentioning that the current analysis considering cerebellar features only yielded a moderate level of prediction accuracy. Including other key brain regions and employing a multimodal approach that integrates different types of brain imaging data may improve the prediction (46,47).

Feature importance analysis for the prediction of SZ highlighted contributions from both motor (48,49) and cognitive regions (15). The limbic vermis, specifically Vermis IX, a region with reported reductions in individuals with SZ (50–52), displayed the highest feature importance in percentage of extreme negative deviations when using the anatomical atlas. This may be interpreted in the context of limbic vermis’s role in emotional processing, facial expression recognition (53,54), and mentalizing, which is integral to understanding others’ mental states, also known as theory of mind (55,56). The involvement of the limbic vermis in these processes is supported by evidence showing connections from this cerebellar region to both cortical and subcortical limbic areas (57). A small study showed smaller inferior posterior lobe in children and adolescents with childhood-onset SZ compared to healthy controls (51), yet, contrasting findings indicate abnormalities may also exist in the anterior lobe (58). A recent study examining a series of 17 individuals with SZ or undifferentiated psychosis (59) showed posterior vermis-predominant cerebellar hypoplasia.

As functional topography does not consistently adhere to anatomical boundaries in the cerebellum, we also examined task-based and resting state atlases. In general, there were both slight discrepancies and shared areas when identifying the features of highest importance across models based on different atlases. Indeed, no one atlas consistently emerged as better than any other. However, we believe that moving between atlases significantly aids in functional interpretation of our findings. For instance, when using the percentage of extreme negative deviations (summarized within ROIs) to predict SZ, cerebellar regions functionally linked to verbal fluency, word comprehension and mental arithmetic (region 9) and limbic A (region 10) exhibited the highest feature importance. And when examining vermis IX in the task-based atlas, it highlights the region of saccades, visual working memory and visual letter recognition (19). Past studies literatures exhibit strong resting-state connectivity between lobules I-VI and vermis VIIB-IX of the cerebellum and the visual network (60), and oculomotor abnormalities are observed in SZ (61). For median, Right I-III showed the highest feature importance in the anatomical regions followed by vermis VIII. On the other hand, a study investigating functional connectivity reported hypoactivation in the vermis III, VI, VII, and VIII, along with a negative correlation between the vermis and time processing abilities in individuals with SZ (62). Moreover, divided attention (region 5) in task-based and Visual B (region 2) and limbic A (region 10) in resting state are highlighted for features in median.

Like SZ (63), ASD is a complex neurodevelopmental condition (12) involving a range of clinical characteristics, including repetitive behaviors, restricted interests, and difficulties in social interaction and communication (64). The substantial heterogeneity in clinical characteristics and severity, ranging from the highest functioning form of autism to those requiring substantial support in their everyday life, makes it challenging to demarcate a common neurobiological underpinning (65). Previous studies did not provide conclusive associations between cerebellar volume and ASD (66). Perhaps related to this, in this study, ASD was significantly associated with variance, i.e. the spread of the deviations within a region. Analyses of feature importance in significant ASD model highlighted Left VIIB and Left Crus II as well as the narrative, emotion and language processing region 7 in the task-based atlas. In addition to median and extreme deviations, variability within cerebellar sub-regions, especially those connected to higher cognitive areas, could thus be a relevant imaging-based marker of ASD. Left VIIB of the anatomical atlas and the region 7 of the task-based atlas overlaps in ASD. These regions of Crus I-II and lobules VIIB are densely connected to the prefrontal and parietal cortices for higher level processes through cerebello-thalamo-cortico-pontine cerebellar circuits (67).

Previous studies have consistently reported good classification of dementia based on imaging data (AUROC ranging from 0.904 to 0.920) (3). Thus, our lack of any significant predictive models for this condition was somewhat surprising (but note that effects of MCI and AD were also relatively small in our previous study (17)). While one must be careful in interpreting null-findings, this lack of any significant effects in a moderately large sample of baseline AD patients nonetheless suggests that the cerebellum is relatively spared (68,69). On the other hand, in both typical aging and AD, grey matter loss in the cerebellum’s Crus I-II and lobule VI is observed, with typical aging showing a bilateral decline and AD in the right hemisphere (70). The cerebellum remains under-studied, and we need to explore how aging and AD pathology contribute to cerebellar atrophy.

There are limitations to consider in our study. First, harmonizing behavioral, cognitive, genetic, phenotypic, lifestyle, symptomatology, and medical history data across various datasets poses significant challenges, especially when aiming for a large sample size essential for assessing generalizability. We assembled a group of participants for whom we had access to essential information such as diagnosis, sex, age, scanning site, and brain imaging data. General limitations in machine learning such as sample size (71) should be taken into consideration when interpreting the current findings. Next, accurately classifying complex clinical conditions is challenging due to the intrinsic heterogeneity of these conditions, which manifests as a wide array of symptoms and genetic variations. Some individuals may exhibit resilience due to genetic or lifestyle factors, which can complicate accurate predictions (72). Further, the existence of sub-groups within heterogeneous conditions, such as ASD, complicates the interpretation of performance metrics of prediction models. Neurodevelopmental changes raise concerns about the appropriateness of applying adult template space and atlases to younger children and adolescents (73). The cerebellum’s distinct position within the skull and its intricate folding pattern also present challenges in obtaining precise MRI data. Finally, an AUROC value in the range of 0.7 to 0.8 can be deemed acceptable for certain clinical applications (74), indicating fair discrimination which includes the range of our model. However, for many clinical scenarios, this may not suffice, as values from 0.8 to 0.9 are generally regarded as appropriate (75). Future research efforts should aim to address these limitations and further enhance our understanding of predictive models.

## Conclusion

This study tested the value of cerebellar-derived features for predictions of five mental and neurological conditions. The analysis revealed moderate prediction performance for ASD and SZ, with strongest contributions from posterior cerebellar regions.

## Data availability

In this study, we used brain imaging from ABIDE, ADNI, AIBL, DEMGEN, and TOP. The cerebellar normative models from this work are available on via PCNportal (76): https://pcnportal.dccn.nl/.

## Code availability

All code used in this work is available at FreeSurfer (https://surfer.nmr.mgh.harvard.edu), FSL (https://fsl.fmrib.ox.ac.uk/fsl/fslwiki/FslInstallation), ACAPULCO (https://gitlab.com/shuohan/acapulco), and SUIT (https://github.com/jdiedrichsen/suit). Code for normative model is available as open-source python package, Predictive Clinical Neuroscience (PCN) toolkit (https://github.com/amarquand/PCNtoolkit). Further code is forked to or published on https://github.com/MHM-lab.

## Ethics of the study

These are analyses of publicly and privately available data. Description of informed consent and other ethical procedures is extensively described in each study, referenced in the manuscript. The data were stored and analyzed using University of Oslo’s secure platform, *Services for sensitive data* (TSD), in compliance with Norwegian privacy regulations.

## Acknowledgements

We are grateful to all the individuals who participated in the studies and acknowledge the contributions of the clinicians and researchers involved in the recruitment and assessment of participants for making this work possible. We want to acknowledge the Norwegian registry of persons assessed for cognitive symptoms (NorCog), for providing access to patient data. We performed this work on the Services for sensitive data (TSD), University of Oslo, Norway, with resources provided by UNINETT Sigma2 - the National Infrastructure for High-Performance Computing and Data Storage in Norway.

Data collection and sharing for this project was funded by the Alzheimer’s Disease Neuroimaging Initiative (ADNI) (National Institutes of Health Grant U01 AG024904) and DOD ADNI (Department of Defense award number W81XWH-12-2-0012). ADNI is funded by the National Institute on Aging, the National Institute of Biomedical Imaging and Bioengineering, and through generous contributions from the following: AbbVie, Alzheimer’s Association; Alzheimer’s Drug Discovery Foundation; Araclon Biotech; BioClinica, Inc.; Biogen; Bristol-Myers Squibb Company; CereSpir, Inc.; Cogstate; Eisai Inc.; Elan Pharmaceuticals, Inc.; Eli Lilly and Company; EuroImmun; F. Hoffmann-La Roche Ltd and its affiliated company Genentech, Inc.; Fujirebio; GE Healthcare; IXICO Ltd.; Janssen Alzheimer Immunotherapy Research & Development, LLC.; Johnson & Johnson Pharmaceutical Research & Development LLC.; Lumosity; Lundbeck; Merck & Co., Inc.; Meso Scale Diagnostics, LLC.; NeuroRx Research; Neurotrack Technologies; Novartis Pharmaceuticals Corporation; Pfizer Inc.; Piramal Imaging; Servier; Takeda Pharmaceutical Company; and Transition Therapeutics. The Canadian Institutes of Health Research is providing funds to support ADNI clinical sites in Canada. Private sector contributions are facilitated by the Foundation for the National Institutes of Health (www.fnih.org). The grantee organization is the Northern California Institute for Research and Education, and the study is coordinated by the Alzheimer’s Therapeutic Research Institute at the University of Southern California. ADNI data are disseminated by the Laboratory for Neuro Imaging at the University of Southern California. Also, data used in preparation of this article were obtained from the Australian Imaging Biomarkers and Lifestyle Study of Ageing (AIBL) databases (adni.loni.usc.edu).

## Authorship Contributions

T.M., T.W., and M.K. originally conceived of the project. M.K., N.S., T.W., T.M., and E.H.L. performed the analyses. M.K. T.W. T.M. wrote the initial draft of the manuscript. O.A., G.R., K.P., D.A., G.S., N.E.S., O.B.S., A.F.M., C.F.B., D.A., T.U., T.W. and L.W. contributed to data curation. All authors discussed the results and contributed to the final manuscript.

