## Supplementary Methods and Tables for "Predicting Mental and Neurological Illnesses Based on Cerebellar Normative Features"

*Table of Contents*

**Kim et al.**

### Supplementary Methods

### List of Supplementary Figures


1. Feature importance of all atlases
2. Between model comparison in anatomical atlas
3. Comparison with Random Forest


### List of Supplementary Tables


1. Sources of the studies used in the study
2. Full sample description and demographics
3. Feature Importance of ASD
4. Feature Importance of BD
5. Feature Importance of SZ
6. Feature Importance of MCI
7. Feature Importance of AD

### Supplementary Methods

#### *Statistics & Reproducibility*

The quality control of this study is identical the procesude from Kim and colleagues(1) utilizing the full sample of the cerebellar normative model. We perfromed quality control by running the ENIGMA Cerebellum Volumetric Pipeline QC Scripts of ACAPULCO(2) with Singularity(3). Given the extensive volume of brain scans available, amounting to thousands, it was not feasible to manually inspect each individual scan. However, approximately 5% of these scans were selected and examined manually to ensure quality and consistency within the dataset. The "QC_Images.html" file displays visual representations of the segmented images in coronal, sagittal, and transverse sections, which allows for a thorough inspection of the segmentation quality. The QC pipeline delivers both quantitative and visual information aids pertaining to the volumetric aspects of the cerebellum's segmented regions including volume, outliers, and box plots of the outliers. By integrating quality control pipeline into our analysis, it enabled us to detect and rectify any mis-segmentations or statistical outliers, thereby enhancing the dependability and precision of our findings. We further excluded participants whose scans revealed outliers in at least two regions, as well as when a scanning site contributed data from fewer than five participants.

#### *Atlases*

The cerebellum's complex structure suggests that its various subregions have distinct roles in different cognitive and motor tasks. Functional subregions within the cerebellum do not correspond neatly with its lobular architecture. As shown in MDTB map(4), specific areas are linked to discrete functions: regions 1 and 2 with hand movements, 3 with visual memory, 4 with attention, 5 and 6 with working memory, 7 and 8 with narrative comprehension, 8 and 9 with language functions, and region 10 with autobiographical recall. These functional delineations show that the organization of cerebellar functions transcends classic anatomical divisions, with functional domains intersecting various lobules.

Buckner et al. (5,6) highlighted the intricate patterns of connectivity between the cerebellum and the cerebral cortex during the resting state. Resting state networks demonstrate shared functional activation despite the absence of explicit tasks. Moereover, cerebellum's participation in continuous functional interactions with the cerebral cortex are crucial for overall brain functionality. The studies have unraveled a comprehensive topographic organization of the cerebellum, with the cerebellar lobules showing specific connectivity patterns with various cortical areas, underpinning the cerebellum's involvement in a spectrum of both motor and non-motor functions. Lobule VIII and the anterior lobe of the cerebellum are connected with cortical and premotor areas that are parts of networks 3, 4, and 7, which are associated with somatomotor and attentional functions. Conversely, lobules VII and IX of the cerebellum are linked with prefrontal and parietal association areas, engaging in networks 8, 12, 13, 14, 16, and 17. The regions for the 17 networks are N1: Visual A, N2: Visual B; Network 3: Somatomotor A; N4: Somatomotor B; N5: Dorsal Attention A; N6: Dorsal Attention B; N7: Salience/ Ventral Attention A; N8: Salience/ Ventral Attention B; N9: Limbic B; N10: Limbic A; N11: Control A; N12: Control B; N13: Control C; N14: Default A; N15: Default B; N16: Default C; N17: Temporal Parietal.

### Supplementary Figures

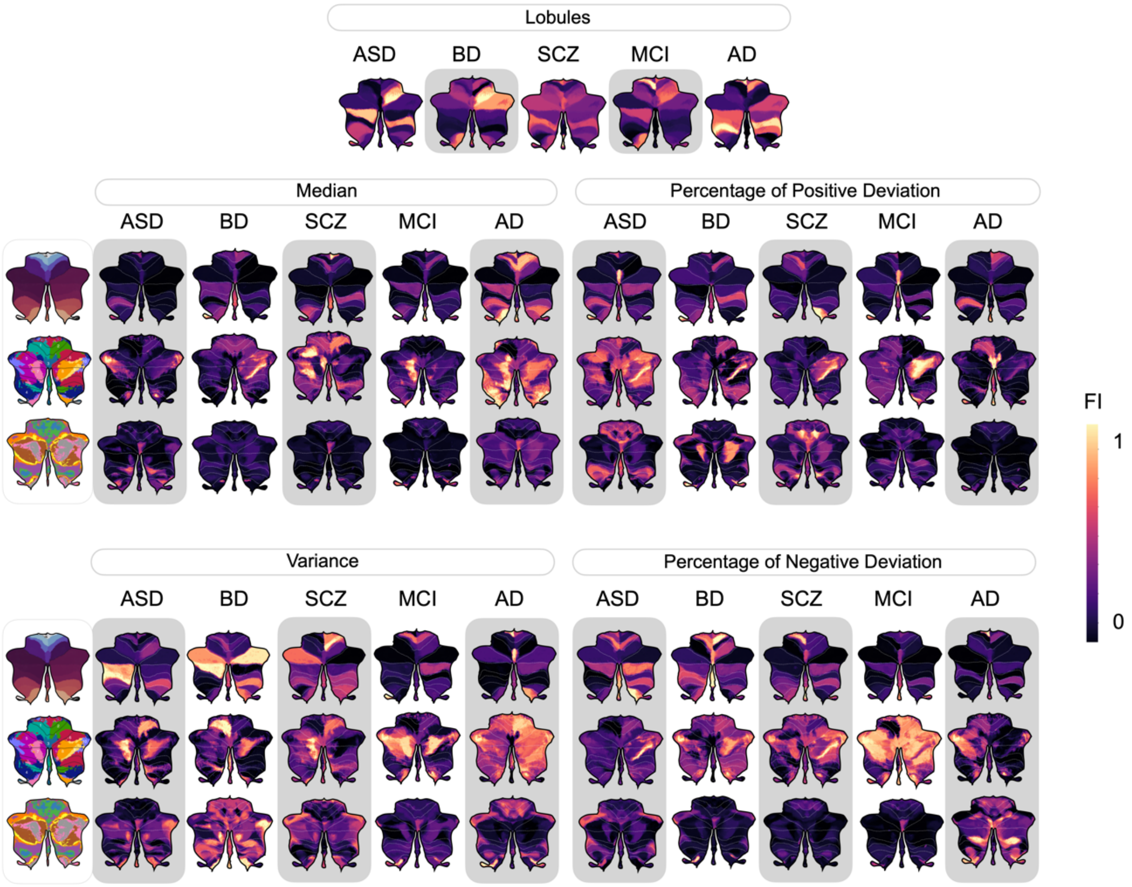


**Supplementary Figure 1. Feature importance of all disorders and atlases shown.** Both features that remained significant and non-significant after adjustments for multiple comparisons of AUROC are shown.

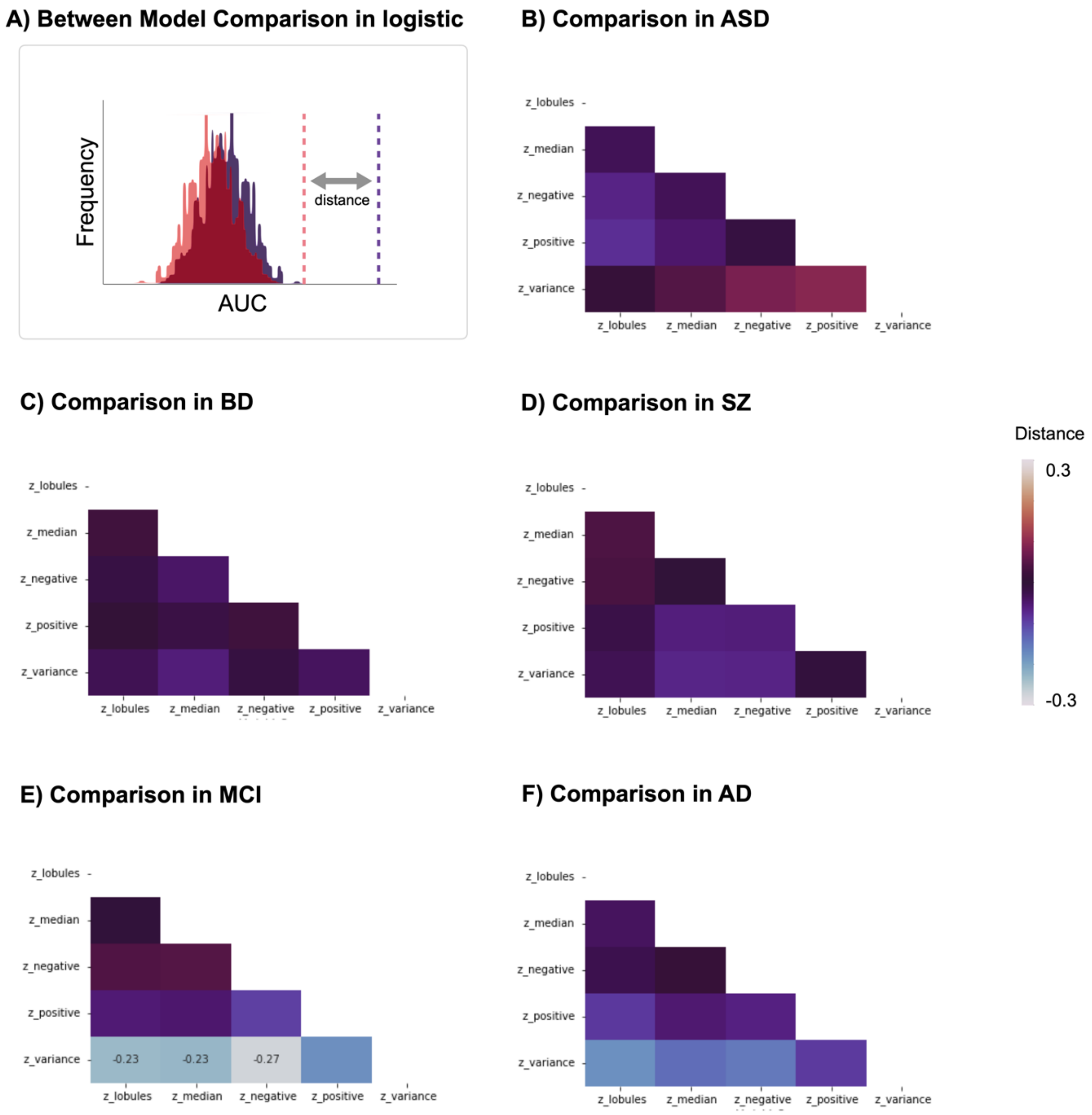


**Supplementary Figure 2. Comparisons between models within the anatomical atlas generally yield similar performance.** (A) The figure illustrates the comparative distance in performance between pairs of models that have withstood multiple comparison tests for each disease across anatomical atlas. (B) This suggests that while most models are equally predictive, the variance model is an exception in the MCI cohort.

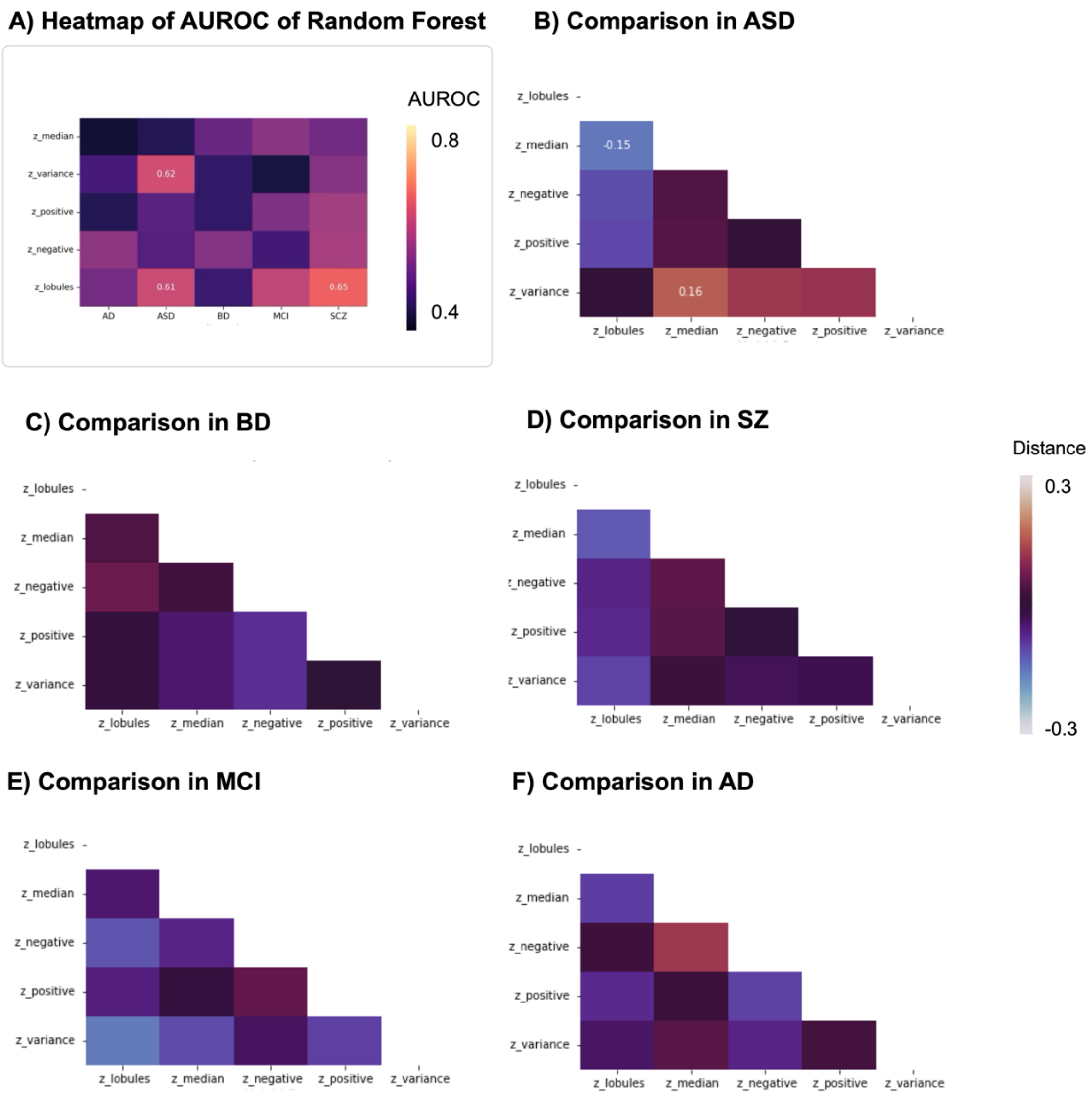


**Supplementary Figure 3. Random Forest achieves a performance comparable to that of Logistic Regressio**n **in anatomical atlas** (A) The heatmap of the AUROC for the Random Forest model displaying results akin to linear regression. (B-F) Also, comparisons between different models using Random Forest also indicating no significant differences except in ASD.

### Supplementary Tables

### Supplementary Table 1. Sources of the studies used in the study

| Datasets | Sources | Comments | References |
|---|---|---|---|
| Autism Brain Imaging Dataset Exchange | http://fcon_1000.projects.nitrc.org/ | Primary support for the work by Adriana Di Martino was provided by the NIMH (K23MH087770) and the Leon Levy Foundation. Primary support for the work by Michael P. Milham and the INDI team was provided by gifts from Joseph P. Healy and the Stavros Niarchos Foundation to the Child Mind Institute, as well as by an NIMH award to MPM (R03MH096321). | (7) |
| Autism Brain Imaging Dataset Exchange II | http://fcon_1000.projects.nitrc.org/ | Primary support for the work by Adriana Di Martino and her team was provided by the National Institute of Mental Health (NIMH 5R21MH107045). Primary support for the work by Michael P. Milham and his team provided by the National Institute of Mental Health (NIMH 5R21MH107045); Nathan S. Kline Institute of Psychiatric Research). Additional Support was provided by gifts from Joseph P. Healey, Phyllis Green and Randolph Cowen to the Child Mind Institute. | (8) |
| Alzheimer's Disease Neuroimaging Initiative | http://adni.loni.usc.edu/ | The ADNI was launched in 2003 as a public-private partnership, led by Principal Investigator Michael W. Weiner, MD. ADNI consists of 4 waves, the later is still ongoing (ADNI 3). A complete listing of ADNI investigators can be found at http://adni.loni.usc.edu/wpcontent/uploads/how_to_apply/ADNI_Acknowledgement_List.pdf. Data collection and sharing for this project was funded by the Alzheimer's Disease Neuroimaging Initiative (ADNI) (National Institutes of Health Grant U01 AG024904) and DOD ADNI (Department of Defense award number W81XWH-12-2-0012). ADNI is funded by the National Institute on Aging, the National Institute of Biomedical Imaging and Bioengineering, and through generous contributions from the following: AbbVie, Alzheimer's Association; Alzheimer's Drug Discovery Foundation; Araclon Biotech; BioClinica, Inc.; Biogen; Bristol-Myers Squibb Company; CereSpir, Inc.; Cogstate; Eisai Inc.; Elan Pharmaceuticals, Inc.; Eli Lilly and Company; EuroImmun; F. Hoffmann-La Roche Ltd and its affiliated company Genentech, Inc.; Fujirebio; GE Healthcare; IXICO Ltd.;Janssen Alzheimer Immunotherapy Research & Development, LLC.; Johnson & Johnson Pharmaceutical Research & Development LLC.; Lumosity; Lundbeck; Merck & Co., Inc.;Meso Scale Diagnostics, LLC.; NeuroRx Research; Neurotrack Technologies; Novartis Pharmaceuticals Corporation; Pfizer Inc.; Piramal Imaging; Servier; Takeda Pharmaceutical Company; and Transition Therapeutics. The Canadian Institutes of Health Research is providing funds to support ADNI clinical sites in Canada. Private sector contributions are facilitated by the Foundation for the National Institutes of Health (www.fnih.org). The grantee organization is the Northern California Institute for Research and Education, and the study is coordinated by the Alzheimer's Therapeutic Research Institute at the University of Southern California. ADNI data are disseminated by the Laboratory for Neuro Imaging at the University of Southern California. | |
| The Australian Imaging, Behaviour and Lifestyle Flagship Study | https://aibl.csiro.au/ | Australian Imaging Biomarkers and Lifestyle flagship study of ageing (AIBL) was funded by the Commonwealth Scientific and Industrial Research Organisation (CSIRO), which was made available at the ADNI databas (http://www.loni.usc.edu/ADNI). The AIBL researchers contributed data but did not participate in analysis or writing of this report. AIBL researchers are listed at http://www.aibl.csiro.au. Correspondence should be addressed to Christopher Rowe. | (9) |
| Demgen | Authors | Supported by the Norwegian National Advisory Unit on Aging and Health | (10,11) |

| | | | |
|---|---|---|---|
| StrokeMRI | Authors | Supported by the Research Council of Norway (249795, 248238), the South-Eastern Norway Regional Health Authority (2014097, 2015044, 2015073, 2016083), and the Norwegian ExtraFoundation for Health and Rehabilitation (2015/FO5146) | (12) |
| TOP | Authors | Supported by several grants from the Research Council of Norway, and the South-Eastern Norway Regional Health Authority | (13–15) |

### Supplementary Table 2. Full Sample description and demographics

| | | N (Participants) | N (Scanning-site) | Age (Mean, S.D.) | Sex (%F:%M) |
|---|---|---|---|---|---|
| Full | All | 54102 | 132 | | |
| | Training set | 27117 | 132 | 54.36 (20.31) | 53:47 |
| | Testing set | 26985 | 132 | 54.52 (20.19) | 53:47 |
| Clinical | Testing set | 1757 | 53 | 29.40 (20.84) | 30:70 |
| | Alzheimer's Disease (AD) | 146 | 13 | 72.42 (7.65) | 53:47 |
| | Autism Spectrum Disorder (ASD) | 900 | 37 | 16.20 (9.00) | 14:86 |
| | Bipolar Disorder (BD) | 277 | 3 | 32.73 (11.67) | 60:40 |
| | Mild Cognitive Impairment (MCI) | 122 | 3 | 67.25 (9.27) | 42:58 |
| | Schizophrenia (SZ) | 312 | 3 | 29.58 (9.52) | 33:67 |

### Supplementary Table 3. Feature Importance of ASD

#### a) Feature Importance of ASD in lobules

| Features | z_lobules_Importance |
|---|---|
| Corpus.Medullare | 0.1743849770904096 |
| Left.Crus.I | 0.1615973502446064 |
| Left.Crus.II | 0.2448591286807397 |
| Left.I.III | 0.0596320262303078 |
| Left.IV | 0.0486371536215285 |
| Left.IX | 0.116279412179556 |
| Left.V | 0.064731872004062 |
| Left.VI | 0.1472285337361797 |
| Left.VIIB | 0.0516654468818476 |
| Left.VIIIA | 0.1272971021375396 |
| Left.VIIIB | 0.1294341201659154 |
| Left.X | 0.2234960365897122 |
| Right.Crus.I | 0.1260286388076124 |
| Right.Crus.II | 0.112025654915322 |
| Right.I.III | 0.0703292426588575 |
| Right.IX | 0.0834227595584971 |
| Right.V | 0.1835404310493908 |
| Right.VI | 0.3647451037469261 |
| Right.VIIB | 0.1626259272812545 |
| Right.VIIIA | 0.0525859748599345 |
| Right.VIIIB | 0.1663374257085011 |
| Right.X | 0.1264992433913717 |
| Rigt.IV | 0.0769340167255874 |
| Vermis.IX | 0.173003371103169 |
| Vermis.VI | 0.0484548634217274 |
| Vermis.VII | 0.0286363152563885 |
| Vermis.VIII | 0.0721832104949135 |
| Vermis.X | 0.2007370242216701 |

#### b) Feature Importance of ASD in anatomical atlas

| Features | z_variance_Importance | z_median_Importance | z_positive_Importance | z_negative_Importance |
|---|---|---|---|---|
| Left.I.III | 0.1751005221454026 | 0.5586269214186221 | 0.4794676392289766 | 0.340714768635482 |
| Right.I.III | 0.5328161128677985 | 0.1390627478515926 | 0.2110546522995292 | 0.1507073858543711 |
| Left.IV | 0.4362305270419605 | 0.1426742507635193 | 0.11994806584145 | 0.2447911384302553 |
| Right.IV | 0.1183853736561793 | 0.2495257324165767 | 0.1431035293800037 | 0.1304371074048695 |
| Left.V | 0.1500952029786246 | 0.2282207916897372 | 0.1086583691566078 | 0.4176796420676735 |
| Right.V | 0.225761365874853 | 0.1028174330265956 | 0.0921210031587837 | 0.3509621188623099 |
| Vermis.VI | 0.318303762365685 | 0.2522114221011555 | 0.7159490460060722 | 0.1359896175719411 |

| | | | | |
|---|---|---|---|---|
| Left.VI | 0.2875527539336978 | 0.0977229258666595 | 0.1450233178241601 | 0.1497090936263107 |
| Right.VI | 0.5399721783059506 | 0.0935764291343389 | 0.0681899818970522 | 0.1325375290705227 |
| Vermis.VII | 0.1531532125353217 | 0.2997250084446862 | 0.4108318832921008 | 0.2105320770694901 |
| Left.Crus.I | 0.2156827005894605 | 0.0756096297164284 | 0.1218314003358821 | 0.1436109904817203 |
| Left.Crus.II | 1.0443443028345003 | 0.1958961880171595 | 0.2993007727674419 | 0.2872748412915805 |
| Left.VIIB | 1.197164811089786 | 0.1334890982403853 | 0.1584582645804186 | 0.0556793411088017 |
| Right.Crus.I | 0.2155102385458175 | 0.0491107722550043 | 0.160848677768186 | 0.0949494596351361 |
| Right.Crus.II | 0.2870190159225087 | 0.0376833736563167 | 0.3822687392005416 | 0.403278506650387 |
| Right.VIIB | 0.3256719581156003 | 0.1292486005697429 | 0.1034695842870924 | 0.1575343193468442 |
| Vermis.VIII | 0.2932396066042603 | 0.1390544755135472 | 0.2480199700172575 | 0.4620168333103452 |
| Left.VIIIA | 0.4526165015576576 | 0.5504454891341128 | 0.4578269722285532 | 0.0874304889230146 |
| Left.VIIIB | 0.348183339795283 | 0.2202777592203245 | 0.18652263191824 | 0.1097562281618798 |
| Right.VIIIA | 0.7450380968504879 | 0.0906474465195491 | 0.155902955261197 | 0.3362888086743448 |
| Right.VIIIIA | 0.5361957882339541 | 0.1129052966110028 | 0.2597223084368795 | 0.168817361713027 |
| Vermis.IX | 0.2939786848284996 | 0.0397337563203992 | 0.4922413076255171 | 0.4624092915901576 |
| Left.IX | 0.1284207224446213 | 0.2373231248211454 | 0.3776167817724774 | 0.2055799867547718 |
| Right.IX | 0.1900668728655543 | 0.195317063571438 | 0.2426803453453687 | 0.5283937722866658 |
| Vermis.X | 0.0637179155212677 | 1.20061999011062 | 0.3451720403367035 | 0.1300139267114345 |
| Left.X | 0.4355807012311487 | 0.6896145675022405 | 0.7478246839784772 | 0.250500760615384 |
| Right.X | 0.3374002204682387 | 0.1141584395399537 | 0.3265623157521213 | 0.3690514014256335 |

### c) Feature Importance of ASD in task-based atlas

| **Features** | **z_variance_Importance** | **z_median_Importance** | **z_positive_Importance** | **z_negative_Importance** |
|---|---|---|---|---|
| 1: Left-hand presses/ motor planning/ interference resolution | 0.1328746872837024 | 0.0293275905523436 | 0.168033176235104 | 0.0427677630046416 |
| 2: Right-hand presses/ motor planning/ divided attention | 0.2776376961878047 | 0.1236045575650287 | 0.0861050121010345 | 0.1967825642751003 |
| 3: Saccades/visual working memory/visual letter recognition | 0.0617585330583789 | 0.0987727571463953 | 0.2112503310334039 | 0.1540437927527996 |
| 4: Action Observation/divided attention/motor planning | 0.0510484548247385 | 0.0292380500065638 | 0.1127222857713337 | 0.1399881698737586 |
| 5: Divided attention/active maintenance/mental arithmetic | 0.1328340103061767 | 0.0865094539918881 | 0.2438754802780273 | 0.1191290761506852 |
| 6: Divided attention/verbal fluency/active maintenance | 0.0617585039359305 | 0.0492285450642826 | 0.2390447840795685 | 0.1077324240935843 |

| | | | | |
|---|---|---|---|---|
| 7: Narrative/ emotion processing/ language processing | 0.3227131620256279 | 0.0703644135588551 | 0.0495938068340027 | 0.0948824028210988 |
| 8: Word comprehension/ language processing/ narrative | 0.2385499293780999 | 0.1226367585293581 | 0.1680882436156004 | 0.1380110613194332 |
| 9: Verbal Fluency/word comprehension/mental arithmetic | 0.1326867904346175 | 0.1643669269176986 | 0.3281880392851926 | 0.418015634627899 |
| 10: Autobiographical recall/visual letter recognition/interference resolution | 0.0510520495325992 | 0.4018787474703671 | 0.0817871938722162 | 0.1838269474588484 |

### d) Feature Importance of ASD in resting-state atlas

| Features | z_variance_Importance | z_median_Importance | z_positive_Importance | z_negative_Importance |
|---|---|---|---|---|
| 1: Visual A | 0.098078935159729 | 0.696307017908821 | 0.1082714068407929 | 0.1375728102963435 |
| 2: Visual B | 0.6535631227083417 | 0.6430107033878375 | 0.7980146900907764 | 0.24663688264155 |
| 3: Somatomotor A | 0.0577498892399181 | 0.0576174782143312 | 0.4791850810549695 | 0.1055868427349992 |
| 4: Somatomotor B | 0.0913366217809954 | 0.1757581862907068 | 0.1069089842715243 | 0.1501324304857504 |
| 5: Dorsal Attention A | 0.6498254136761844 | 0.2173284350460524 | 0.7881573297427298 | 0.7059790608127074 |
| 6: Dorsal Attention B | 0.3597909703243311 | 0.4841860908098586 | 0.2423922667271315 | 0.0687138605883437 |
| 7: Salience/Ventral Attention A | 0.1140707704963555 | 0.0764909457263593 | 0.560016853434153 | 0.2671888482345047 |
| 8: Salience/ Ventral Attention B | 0.1476531611130337 | 0.1388648151455638 | 0.1095928450167874 | 0.4833919085765815 |
| 9: Limbic B | 0.1726738083921067 | 0.1333127971646131 | 0.1108769208552548 | 0.2037632856482712 |
| 10: Limbic A | 0.1296903070145517 | 0.19520243855016 | 0.7279348656615604 | 0.1970761940846492 |
| 11: Control A | 0.103421481256824 | 0.0839770891773178 | 0.1671481640654739 | 0.4474436624244468 |
| 12 Control B | 0.0577498892604256 | 0.0687579112555635 | 0.1737117193671591 | 0.0790458757091334 |
| 13: Control C | 0.2482220918090958 | 0.0276659767477823 | 0.131641589371915 | 0.0704257916400572 |
| 14: Default A | 0.2892020323726066 | 0.1813819134938358 | 0.263985594236554 | 0.0764372346125098 |
| 15: Default B | 0.193442683787445 | 0.3135701931225153 | 0.2362953785654634 | 0.2004281049150354 |
| 16: Default C | 0.5065933514592946 | 0.2634700834053719 | 0.0723407812452423 | 0.0343677938163116 |
| 17: Temporal Parietal | 0.2482616408769188 | 0.0447636546126759 | 0.2543980628836862 | 0.2025824928976594 |

### Supplementary Table 4. Feature Importance of BD

#### a) Feature Importance of BD in lobules

| Features | z_lobules_Importance |
|---|---|
| Corpus.Medullare | 0.1799552246335557 |
| Left.Crus.I | 0.080817930156383 |
| Left.Crus.II | 0.0842803480357426 |
| Left.I.III | 0.1151190596353922 |
| Left.IV | 0.1062222733138265 |
| Left.IX | 0.3064084844756838 |
| Left.V | 0.0990674949665181 |
| Left.VI | 0.196343571556421 |
| Left.VIIB | 0.066931910130356 |
| Left.VIIIA | 0.033557889908478 |
| Left.VIIIB | 0.0854289387915874 |
| Left.X | 0.1569906088828636 |
| Right.Crus.I | 0.2221395206629539 |
| Right.Crus.II | 0.1519249420279759 |
| Right.I.III | 0.1089381667099158 |
| Right.IX | 0.2155438149495519 |
| Right.V | 0.0335707613193793 |
| Right.VI | 0.3517208547631265 |
| Right.VIIB | 0.0669441085928049 |
| Right.VIIIA | 0.0675645205441756 |
| Right.VIIIB | 0.1083282131509523 |
| Right.X | 0.1330985841266789 |
| Rigt.IV | 0.1877757610222981 |
| Vermis.IX | 0.2393580380609084 |
| Vermis.VI | 0.1872539200969538 |
| Vermis.VII | 0.14795246679034 |
| Vermis.VIII | 0.0976054370887731 |
| Vermis.X | 0.0458760969799567 |

#### b) Feature Importance of BD in anatomical

| Features | z_variance_Importance | z_median_Importance | z_positive_Importance | z_negative_Importance |
|---|---|---|---|---|
| Left.I.III | 0.1920800064794396 | 0.2234278383435405 | 0.378477362019497 | 0.306108802885538 |
| Right.I.III | 0.2044663758953064 | 0.3915107702409685 | 0.1475602505889362 | 0.3955028023028737 |
| Left.IV | 0.2854434652155219 | 0.2560918078019666 | 0.1726806455554318 | 0.4749947902355365 |
| Right.IV | 0.19762422982742 | 0.3387179886279116 | 0.1042438018511475 | 0.6380422879496778 |
| Left.V | 0.4115413717735392 | 0.2334564607998918 | 0.1612775682044664 | 0.5210858798477698 |
| Right.V | 0.2580946485660995 | 0.0773265212572336 | 0.4677620821148521 | 0.1333398989777899 |
| Vermis.VI | 0.3245719635771267 | 0.2558678631690986 | 0.1238174122641022 | 0.5398555667383652 |

| | | | | |
|---|---|---|---|---|
| Left.VI | 0.194381311580095 | 0.1034735164542756 | 0.1968194144641407 | 0.0919341427765402 |
| Right.VI | 0.1986345042733759 | 0.050792420185594 | 0.160358701883074 | 0.3741404376201672 |
| Vermis.VII | 0.4428898361410314 | 0.3481989550402251 | 0.4312485212746445 | 0.6105738212947909 |
| Left.Crus.I | 0.604923800122313 | 0.0914910541650948 | 0.2248113382558756 | 0.1829690290685078 |
| Left.Crus.II | 0.7501588196521196 | 0.2817317266470223 | 0.0491079196589898 | 0.2487564379753132 |
| Left.VIIB | 0.1608881264119955 | 0.2707228815667318 | 0.1696528326964148 | 0.4240268854442321 |
| Right.Crus.I | 0.7209618045066964 | 0.0535544473335531 | 0.1352709222649734 | 0.1215253465271902 |
| Right.Crus.II | 0.0800622896954081 | 0.0832409024488672 | 0.2154883953188147 | 0.2955814089353234 |
| Right.VIIB | 0.2812442375879079 | 0.2007359154718721 | 0.5289453867559091 | 0.2393138616232433 |
| Vermis.VIII | 0.4700930895451081 | 0.4816618527675134 | 0.4453220955263516 | 0.6310573368785376 |
| Left.VIIIA | 0.1115439572841212 | 0.0554950839792504 | 0.2558729762039585 | 0.3422335837760663 |
| Left.VIIIB | 0.0800524654974194 | 0.2630411739743976 | 0.278816501596459 | 0.1544891962996259 |
| Right.VIIIA | 0.5925698349672184 | 0.1211202861387443 | 0.2628752863883717 | 0.1916438874105798 |
| Right.VIIIIA | 0.2711403595704053 | 0.2153962721853383 | 0.1064472889605652 | 0.3230974397272569 |
| Vermis.IX | 0.1537576372671153 | 0.3522991155909024 | 0.245414574410637 | 0.2017647350169163 |
| Left.IX | 0.2934879235678734 | 0.3624499859059373 | 0.3810063900717059 | 0.3907452637839671 |
| Right.IX | 0.5840394100659877 | 0.0924610940102935 | 0.1250739350577188 | 0.1206763684550131 |
| Vermis.X | 0.3501067643369872 | 0.2365767114756475 | 0.4705229184040426 | 0.2843653776568503 |
| Left.X | 0.1825451420874197 | 0.734499625104544 | 0.8395759454285742 | 0.1919160180606601 |
| Right.X | 0.3949037636273318 | 0.2114974649199087 | 0.2122033349384194 | 0.3893217142950423 |

### c) Feature Importance of BD in task-based

| Features | z_variance_Importance | z_median_Importance | z_positive_Importance | z_negative_Importance |
|---|---|---|---|---|
| 1: Left-hand presses/ motor planning/ interference resolution | 1.131264485542982 | 0.0378874453769629 | 0.1233298563876228 | 0.1781998041075252 |
| 2: Right-hand presses/ motor planning/ divided attention | 0.0525476339859855 | 0.1038545166952326 | 0.1518885697913079 | 0.2433956408177569 |
| 3: Saccades/visual working memory/visual letter recognition | 0.6225637029918147 | 0.1169171148883951 | 0.1456759252892859 | 0.0930957895774605 |
| 4: Action Observation/divided attention/motor planning | 0.0959113381041849 | 0.0375348003380781 | 0.1884092812137374 | 0.2173032409422664 |
| 5: Divided attention/active maintenance/mental arithmetic | 0.1006891828539643 | 0.0648484520510779 | 0.089676410253079 | 0.1018464771663426 |
| 6: Divided attention/verbal fluency/active maintenance | 0.4860951554894037 | 0.0931767376175608 | 0.0650801045510374 | 0.2032556650346712 |

| | | | | |
|---|---|---|---|---|
| 7: Narrative/ emotion processing/ language processing | 0.0524127328303683 | 0.0713578163415512 | 0.2025109148095158 | 0.1297586227947881 |
| 8: Word comprehension/ language processing/ narrative | 0.0439241543998877 | 0.1123011517555026 | 0.1340654843540411 | 0.3285723608012487 |
| 9: Verbal Fluency/word comprehension/mental arithmetic | 0.0495004677251966 | 0.2227742476291548 | 0.276179719425365 | 0.1661271557190477 |
| 10: Autobiographical recall/visual letter recognition/interference resolution | 0.0959113467381921 | 0.0494098816399451 | 0.123373075328259 | 0.3800585876243454 |

### d) Feature Importance of BD in resting-state

| Features | z_variance_Importance | z_median_Importance | z_positive_Importance | z_negative_Importance |
|---|---|---|---|---|
| 1: Visual A | 0.048223071568384 | 0.5338083924278971 | 0.1838734008331006 | 0.3091412893409924 |
| 2: Visual B | 0.1793396121930982 | 0.1699157360865917 | 0.3452417921238152 | 0.8021607219410057 |
| 3: Somatomotor A | 0.4835731303350169 | 0.2611137850575308 | 0.1057660959829964 | 0.1408272606179943 |
| 4: Somatomotor B | 0.0120963693760497 | 0.1081585415848229 | 0.0906014587199315 | 0.2333499652559711 |
| 5: Dorsal Attention A | 0.2010468966455373 | 0.3835444794864686 | 0.2746583113269051 | 0.1438094658459718 |
| 6: Dorsal Attention B | 0.2019482036345819 | 0.1165062029149344 | 0.1071492026534121 | 0.1343966554407055 |
| 7: Salience/Ventral Attention A | 0.5735510571277656 | 0.1272990064545163 | 0.0699859855707002 | 0.1978864885257224 |
| 8: Salience/ Ventral Attention B | 0.2044322052802461 | 0.2526330196320033 | 0.3375230720162676 | 0.3789291298330217 |
| 9: Limbic B | 0.4923124161523584 | 0.1150217046375819 | 0.149491775723891 | 0.3347925117247042 |
| 10: Limbic A | 0.2776798433860677 | 1.1643548535941146 | 0.149778387855514 | 0.2801945950263801 |
| 11: Control A | 0.1224120990114089 | 0.1034934590878609 | 0.0839041099321614 | 1.0447769738234878 |
| 12 Control B | 0.6965711088698067 | 0.2979520147549359 | 0.0799486360595698 | 0.073100852819005 |
| 13: Control C | 0.0956212698522669 | 0.1579966339581245 | 0.0421802639214098 | 0.0670995724711147 |
| 14: Default A | 0.1995861613776418 | 0.3094807230570077 | 0.4512731643958596 | 0.2527295882774893 |
| 15: Default B | 0.2186406366337339 | 0.1368300979907229 | 0.6176651965969834 | 0.1660509892285661 |
| 16: Default C | 0.8814196036391648 | 0.0957290088940107 | 0.1806881171024322 | 0.2217434212596816 |
| 17: Temporal Parietal | 0.3614628843911893 | 0.3165360528361685 | 0.4958629116913644 | 0.2635990557828341 |

**Supplementary Table 5. Feature Importance of SZ**

a) **Feature Importance of SZ in lobules**

| Features | z_lobules_Importance |
|---|---|
| Corpus.Medullare | 0.0929983713466658 |
| Left.Crus.I | 0.2764924757100045 |
| Left.Crus.II | 0.0741047657753078 |
| Left.I.III | 0.3232797279555975 |
| Left.IV | 0.1671425781557969 |
| Left.IX | 0.1083299745698336 |
| Left.V | 0.1102896951082006 |
| Left.VI | 0.2073714528890709 |
| Left.VIIB | 0.2599524462058514 |
| Left.VIIIA | 0.059276720202208 |
| Left.VIIIB | 0.0996909696897221 |
| Left.X | 0.0907345144831736 |
| Right.Crus.I | 0.0370951289412171 |
| Right.Crus.II | 0.1249554077839712 |
| Right.I.III | 0.1857378812487713 |
| Right.IX | 0.2391041868602232 |
| Right.V | 0.0951724923214643 |
| Right.VI | 0.21596267290435 |
| Right.VIIB | 0.2682841257806463 |
| Right.VIIIA | 0.0756170291511343 |
| Right.VIIIB | 0.1220156379626365 |
| Right.X | 0.0550208719968741 |
| Rigt.IV | 0.0796743563710287 |
| Vermis.IX | 0.2719326982043935 |
| Vermis.VI | 0.1390564927178177 |
| Vermis.VII | 0.1247276509193895 |
| Vermis.VIII | 0.0828191713601032 |
| Vermis.X | 0.166432052560991 |

b) **Feature Importance of SZ in anatomical atlas**

| Features | z_variance_Importance | z_median_Importance | z_positive_Importance | z_negative_Importance |
|---|---|---|---|---|
| Left.I.III | 0.2027224241800506 | 0.2447497560236897 | 0.3847113044899385 | 0.2213698073195407 |
| Right.I.III | 0.7085808727449422 | 0.5671775126197754 | 0.2471613329380528 | 0.2096960046816671 |
| Left.IV | 0.0673210171171232 | 0.0565568078165377 | 0.2485821251926917 | 0.5180900628561951 |
| Right.IV | 0.6674245471741781 | 0.0927031763723532 | 0.1570188612899637 | 0.1702642466297228 |
| Left.V | 0.1765528304906803 | 0.276430203695488 | 0.6011200523637947 | 0.1916224178937363 |
| Right.V | 0.8714145932520854 | 0.1801932880423385 | 0.3069387703289858 | 0.1798920605341969 |
| Vermis.VI | 0.3178710367744907 | 0.1092010693096138 | 0.3268583474183106 | 0.3455736070370718 |

| | | | | |
|---|---|---|---|---|
| Left.VI | 0.1639683963690461 | 0.0660678873741362 | 0.3107413805918132 | 0.0606090934308043 |
| Right.VI | 0.2821274732873535 | 0.0856141579595015 | 0.0903312714290664 | 0.1640425057751334 |
| Vermis.VII | 0.3369378427563906 | 0.1279570053253199 | 0.1556134772891727 | 0.0686525913777373 |
| Left.Crus.I | 0.6227515737403613 | 0.0671808396890814 | 0.1144661217580341 | 0.0978715722349811 |
| Left.Crus.II | 0.2616436244810839 | 0.0235263420106807 | 0.0539817839341485 | 0.0991928668072611 |
| Left.VIIB | 0.3235658869199757 | 0.0541876381351496 | 0.246216882740107 | 0.1544538200908974 |
| Right.Crus.I | 0.0599713602576175 | 0.0238533938858206 | 0.116080687707285 | 0.0584611640870991 |
| Right.Crus.II | 0.2480801017746028 | 0.0626161032875024 | 0.1132042204216976 | 0.2702996904794937 |
| Right.VIIB | 0.4574337801956227 | 0.2925785918174148 | 0.1639444772087586 | 0.127118431505273 |
| Vermis.VIII | 0.506635516080679 | 0.4128747811910787 | 0.2429794501494419 | 0.292193226713242 |
| Left.VIIIA | 0.4593816494137738 | 0.1131033535977099 | 0.2079173279140334 | 0.2256131758373126 |
| Left.VIIIB | 0.0673210171170141 | 0.3541559622596695 | 0.2476827680008449 | 0.2577537473557069 |
| Right.VIIIA | 0.426187859958962 | 0.1269936889914227 | 0.2891487013565553 | 0.3066090448390963 |
| Right.VIIIIA | 0.5289693031197369 | 0.0649811901462652 | 0.2139879037090732 | 0.1633684081763333 |
| Vermis.IX | 0.2210019834768806 | 0.0933803239164193 | 0.4164919738191576 | 0.5593403128706865 |
| Left.IX | 0.4593816484093838 | 0.1565192432648791 | 0.1752919501353453 | 0.2997170188340455 |
| Right.IX | 0.4753733289041975 | 0.0631999474163569 | 0.8957772149712848 | 0.1130630431684946 |
| Vermis.X | 0.4375328395466092 | 0.1739784100851624 | 0.3112178137368669 | 0.2161084518550182 |
| Left.X | 0.3793789181098635 | 0.2788366568989424 | 0.5080133251111405 | 0.163142602518287 |
| Right.X | 0.0997330611555012 | 0.2917535657881785 | 0.6492796882339581 | 0.4887150056873762 |

### c) Feature Importance of SZ in task based atlas

| Features | z_variance_Importance | z_median_Importance | z_positive_Importance | z_negative_Importance |
|---|---|---|---|---|
| 1: Left-hand presses/ motor planning/ interference resolution | 0.1720696089892944 | 0.08919703215821 | 0.1152782930480698 | 0.1055516153181767 |
| 2: Right-hand presses/ motor planning/ divided attention | 0.8041732577313512 | 0.1033415897977294 | 0.0513245874132623 | 0.0775643976148585 |
| 3: Saccades/visual working memory/visual letter recognition | 0.6107795080063653 | 0.0655503944706932 | 0.1160572242953406 | 0.1318407596171336 |
| 4: Action Observation/divided attention/motor planning | 0.3896394055408999 | 0.0772451309081957 | 0.1414463349263642 | 0.0404920883656944 |
| 5: Divided attention/active maintenance/mental arithmetic | 0.6107795080112406 | 0.1207284523951974 | 0.068430266922408 | 0.0511896025696084 |
| 6: Divided attention/verbal fluency/active maintenance | 0.38963941315708 | 0.0570446614772989 | 0.0877486853821974 | 0.1337997389425212 |

| | | | | |
|---|---|---|---|---|
| 7: Narrative/ emotion processing/ language processing | 1.0804509836785905 | 0.0616351250280552 | 0.1201243610638954 | 0.1394917811264521 |
| 8: Word comprehension/ language processing/ narrative | 0.0980479799870637 | 0.0751325445956171 | 0.2237758770692164 | 0.0977187396696492 |
| 9: Verbal Fluency/word comprehension/mental arithmetic | 0.1395219987533257 | 0.0531575012277929 | 0.3953020457741966 | 0.2045383932503965 |
| 10: Autobiographical recall/visual letter recognition/interference resolution | 0.1395220001328581 | 0.0537159705010237 | 0.1447261297522597 | 0.1860810009763548 |

### d) Feature Importance of SZ in resting-state atlas

| Features | z_variance_Importance | z_median_Importance | z_positive_Importance | z_negative_Importance |
|---|---|---|---|---|
| 1: Visual A | 0.2064994235755196 | 0.1344372766603844 | 0.1761139435981971 | 0.2345815575495018 |
| 2: Visual B | 0.5126178013019935 | 0.5826234545716603 | 0.4569628080484274 | 0.240000646261518 |
| 3: Somatomotor A | 0.6335914522860427 | 0.1171298739689782 | 0.0999241519176964 | 0.5123855963031897 |
| 4: Somatomotor B | 0.981733176001331 | 0.0890774149972657 | 0.4582036136183703 | 0.2829495993918292 |
| 5: Dorsal Attention A | 0.2285674054599447 | 0.2120321374291632 | 0.4304223720265486 | 1.3388088294962937 |
| 6: Dorsal Attention B | 0.3894418544113063 | 0.059964028690854 | 0.0351939525262716 | 0.2705040381299158 |
| 7: Salience/Ventral Attention A | 0.5481822792829105 | 0.0311985373370047 | 0.2459497995991687 | 0.2478122056267497 |
| 8: Salience/ Ventral Attention B | 0.7630793371142073 | 0.0737420389545523 | 0.1005284222536441 | 0.1014115234105279 |
| 9: Limbic B | 0.5488813698093589 | 0.1801472616315846 | 0.2825999677719701 | 0.1211046604125484 |
| 10: Limbic A | 0.4794392635634604 | 0.4837188381972558 | 0.2523598459442756 | 0.6888551737799062 |
| 11: Control A | 0.6541395805543945 | 0.1209674107209827 | 0.1264761042220956 | 0.1463022993296994 |
| 12 Control B | 0.9123871863011188 | 0.0358171513879105 | 0.1185045632796862 | 0.1871745735611745 |
| 13: Control C | 0.0382716853131812 | 0.0774365849035534 | 0.0790324525117317 | 0.2970585162672144 |
| 14: Default A | 0.1713020720924531 | 0.2726385121085264 | 0.123410294052409 | 0.3826169895494855 |
| 15: Default B | 0.3580660649407394 | 0.2196943432904239 | 0.2638808179688721 | 0.235327268679623 |
| 16: Default C | 0.038271685183955 | 0.1012152705032945 | 0.0662870783777236 | 0.1721877153073197 |
| 17: Temporal Parietal | 0.555673649312937 | 0.0528628479397919 | 0.2149450569117799 | 0.103366899708517 |

### Supplementary Table 6. Feature Importance of MCI

#### a) Feature Importance of MCI in lobular

| Features | z_lobules_Importance |
|---|---|
| Corpus.Medullare | 0.4386693714066485 |
| Left.Crus.I | 0.1366428680413801 |
| Left.Crus.II | 0.3975531792331915 |
| Left.I.III | 0.2228342196566111 |
| Left.IV | 0.6650713061510655 |
| Left.IX | 0.5234242274914058 |
| Left.V | 0.1504949181066108 |
| Left.VI | 0.2989744349142021 |
| Left.VIIB | 0.2149009895514267 |
| Left.VIIIA | 0.1126313898458978 |
| Left.VIIIB | 0.0664532320689508 |
| Left.X | 0.0998360680758911 |
| Right.Crus.I | 0.2581351234246005 |
| Right.Crus.II | 0.1744007731567407 |
| Right.I.III | 0.4648801534780936 |
| Right.IX | 0.2169992008204193 |
| Right.V | 0.4214077664555102 |
| Right.VI | 0.4923461166248692 |
| Right.VIIB | 0.1373316985681268 |
| Right.VIIIA | 0.1481128179974782 |
| Right.VIIIB | 0.1776206938944213 |
| Right.X | 0.1609113667013718 |
| Rigt.IV | 0.2492917271582767 |
| Vermis.IX | 0.1044793733766818 |
| Vermis.VI | 0.2734046427366169 |
| Vermis.VII | 0.0556361865898493 |
| Vermis.VIII | 0.0635396306844705 |
| Vermis.X | 0.1167290611077794 |

#### b) Feature Importance of MCI in anatomical

| Features | z_variance_Importance | z_median_Importance | z_positive_Importance | z_negative_Importance |
|---|---|---|---|---|
| Left.I.III | 0.0661968393956307 | 0.0664047333614159 | 0.2585308776319255 | 0.2023978948031591 |
| Right.I.III | 0.1722406898604378 | 0.1654064014055538 | 0.163192244387604 | 0.4474610642320016 |
| Left.IV | 0.4018369822543394 | 0.0493351808298168 | 0.1282751330835837 | 0.2922485481868864 |
| Right.IV | 0.3248554958008973 | 0.0916320161702711 | 0.0923320470032784 | 0.3014251432070541 |
| Left.V | 0.176021485715931 | 0.1219258324363601 | 0.1286395249805779 | 0.1603746948164778 |
| Right.V | 0.1992535360293723 | 0.1980090358266978 | 0.1080108521662352 | 0.0892361587365141 |

| | | | | |
|---|---|---|---|---|
| Vermis.VI | 0.1388549569324273 | 0.1343497521304072 | 0.5160547825010615 | 0.4323264878953389 |
| Left.VI | 0.0661968393982973 | 0.0342860227697314 | 0.0757616045748413 | 0.0657068179814799 |
| Right.VI | 0.1822032217027083 | 0.0540362125777007 | 0.1161053570556537 | 0.070428785747495 |
| Vermis.VII | 0.1389613584021006 | 0.1183489684339095 | 0.6047144917001499 | 0.4785033073873074 |
| Left.Crus.I | 0.0690056090849357 | 0.0498402972766119 | 0.1468901870011613 | 0.0196847239900093 |
| Left.Crus.II | 0.1181312424834386 | 0.1684620086930175 | 0.164966005647094 | 0.3059787575938867 |
| Left.VIIB | 0.1388549578021596 | 0.0440860797593695 | 0.0814997382165892 | 0.0401337624573971 |
| Right.Crus.I | 0.0661054841684845 | 0.0312698345497241 | 0.0801872125702411 | 0.0317944349793526 |
| Right.Crus.II | 0.3007160883591536 | 0.0478033400148611 | 0.0865758569721154 | 0.1400366457579256 |
| Right.VIIB | 0.0600614449750787 | 0.1266783927086682 | 0.2219394879235576 | 0.0892447479926076 |
| Vermis.VIII | 0.1751953912327249 | 0.1198206591828881 | 0.3630899159889151 | 0.5427304308072383 |
| Left.VIIIA | 0.0974282948404972 | 0.063102446551449 | 0.080209142005057 | 0.0838526945193687 |
| Left.VIIIB | 0.1528817837113703 | 0.1849740991458323 | 0.1083426672905461 | 0.1326871626082505 |
| Right.VIIIA | 0.1605564751558874 | 0.0876486233545408 | 0.0768112596659515 | 0.2361480013871923 |
| Right.VIIIIA | 0.1680984232901763 | 0.0980396166488705 | 0.2316388950903936 | 0.0680216019642153 |
| Vermis.IX | 0.2270238808584099 | 0.4025528132903853 | 0.1699298111316896 | 0.7712056741490168 |
| Left.IX | 0.0818420921662485 | 0.0960127692413241 | 0.1192380600011127 | 0.1862070537035936 |
| Right.IX | 0.1764484985032052 | 0.0980595562075191 | 0.1687492321839773 | 0.2069058259977817 |
| Vermis.X | 0.1906492972595933 | 0.1103530612376814 | 0.2015574643597655 | 0.6198252235045365 |
| Left.X | 0.5341041556428029 | 0.1976045306429153 | 0.4704546346653602 | 0.3795296479670866 |
| Right.X | 0.2093311480975858 | 0.2930105783145543 | 0.3811206065794021 | 0.9866493036043 |

### c) Feature Importance of MCI in task-based

| Features | z_variance_Importance | z_median_Importance | z_positive_Importance | z_negative_Importance |
|---|---|---|---|---|
| 1: Left-hand presses/ motor planning/ interference resolution | 0.1438158539048763 | 0.0372386664801141 | 0.1096879852523285 | 0.0316237295357179 |
| 2: Right-hand presses/ motor planning/ divided attention | 0.2142848478762932 | 0.023816919498807 | 0.0503179387396081 | 0.2054090054287737 |
| 3: Saccades/visual working memory/visual letter recognition | 0.2453033911893838 | 0.1023024260089984 | 0.0942427344933 | 0.1396480742977144 |
| 4: Action Observation/divided attention/motor planning | 0.1400624954166904 | 0.082017334175404 | 0.1310263373640978 | 0.0486257369554451 |
| 5: Divided attention/active maintenance/mental arithmetic | 0.4591986958517654 | 0.0696941246849814 | 0.074024190895449 | 0.1815314085142669 |
| 6: Divided attention/verbal fluency/active maintenance | 0.2176191869522864 | 0.0580596901826189 | 0.0634333081862864 | 0.2398593257047916 |

| | | | | |
|---|---|---|---|---|
| 7: Narrative/ emotion processing/ language processing | 0.3736089986117381 | 0.1781793983114841 | 0.0763110219180305 | 0.1522312697006042 |
| 8: Word comprehension/ language processing/ narrative | 0.3925349192140037 | 0.0710163610733933 | 0.2719017309779599 | 0.177059553224447 |
| 9: Verbal Fluency/word comprehension/mental arithmetic | 0.1438158601201758 | 0.102767116410945 | 0.2123837733128014 | 0.0687332259849522 |
| 10: Autobiographical recall/visual letter recognition/interference resolution | 0.44145331989946 | 0.0505104861590716 | 0.1200635087639405 | 0.2521567001031988 |

### d) Feature Importance of MCI in resting-state

| Features | z_variance_Importance | z_median_Importance | z_positive_Importance | z_negative_Importance |
|---|---|---|---|---|
| 1: Visual A | 0.0675852989673975 | 0.1505940079018162 | 0.1397209652409215 | 0.2693563045016747 |
| 2: Visual B | 0.4632844876443888 | 0.1393923351600078 | 0.2494283933304406 | 0.7538914700225245 |
| 3: Somatomotor A | 0.1214267524821259 | 0.1295400813391087 | 0.0507308383733983 | 0.1217252047070787 |
| 4: Somatomotor B | 0.1214267524914149 | 0.1662245995825844 | 0.2351788796666568 | 0.0841626311307977 |
| 5: Dorsal Attention A | 0.4080829729076882 | 0.2943080166327639 | 0.6145210469229396 | 0.1412515535539789 |
| 6: Dorsal Attention B | 0.2124458958658857 | 0.0354898525728809 | 0.1196454182851353 | 0.1016460344596897 |
| 7: Salience/Ventral Attention A | 0.1941377732314179 | 0.0247554066210296 | 0.1705232198510103 | 0.1215479453706828 |
| 8: Salience/ Ventral Attention B | 0.0827314169172363 | 0.0445803925543806 | 0.0797993728947138 | 0.0213175715988497 |
| 9: Limbic B | 0.3575223644673026 | 0.1386996356787022 | 0.1290200169810238 | 0.3220415501240724 |
| 10: Limbic A | 0.2654713995116996 | 0.4905222196966898 | 0.2299097883171779 | 0.226957602152739 |
| 11: Control A | 0.6863956075786557 | 0.0671021523782984 | 0.2391207716825845 | 0.1911297256065433 |
| 12 Control B | 0.222187717311119 | 0.0789788892813627 | 0.2526945748068033 | 0.07884875209607 |
| 13: Control C | 0.0599090311500504 | 0.0545829151514611 | 0.0512631427078581 | 0.0866970606402684 |
| 14: Default A | 0.1680355891987581 | 0.8383450735319743 | 0.2631931200230131 | 0.7778030918130132 |
| 15: Default B | 0.3087678144393335 | 0.4281614798083078 | 0.1655807932326203 | 0.5786965936375856 |
| 16: Default C | 0.2124458955701016 | 0.0647653123867089 | 0.1564771467862746 | 0.0797669619466831 |
| 17: Temporal Parietal | 0.2124458954710265 | 0.0792167630065341 | 0.062946064680553 | 0.0662614129819923 |

### Supplementary Table 7. Feature Importance of AD

#### a) Feature Importance of AD in lobular

| Features | z_lobules_Importance |
|---|---|
| Corpus.Medullare | 0.2799181596910814 |
| Left.Crus.I | 0.156152406030598 |
| Left.Crus.II | 0.3492485991097134 |
| Left.I.III | 0.4888405758926621 |
| Left.IV | 0.08946323365111 |
| Left.IX | 0.1328984951667997 |
| Left.V | 0.0919700271057945 |
| Left.VI | 0.1806987309471607 |
| Left.VIIB | 0.3666986042602335 |
| Left.VIIIA | 0.5034017040919078 |
| Left.VIIIB | 0.1959306650940625 |
| Left.X | 0.1749992602935006 |
| Right.Crus.I | 0.3154955322553164 |
| Right.Crus.II | 0.228769972250887 |
| Right.I.III | 0.2809906869848318 |
| Right.IX | 0.1524606476241743 |
| Right.V | 0.158619631226495 |
| Right.VI | 0.1088270377514275 |
| Right.VIIB | 0.4810509558424178 |
| Right.VIIIA | 0.3589520029587559 |
| Right.VIIIB | 0.0696972571656974 |
| Right.X | 0.1374303340614648 |
| Rigt.IV | 0.4170114228949842 |
| Vermis.IX | 0.0794542226196939 |
| Vermis.VI | 0.1345488364540123 |
| Vermis.VII | 0.1228289958278003 |
| Vermis.VIII | 0.0732037146652119 |
| Vermis.X | 0.2764488148887988 |

#### b) Feature Importance of AD in anatomical atlas

| Features | z_variance_Importance | z_median_Importance | z_positive_Importance | z_negative_Importance |
|---|---|---|---|---|
| Left.I.III | 0.8267289050836932 | 0.2039016971725526 | 0.1000003524885001 | 1.0486898580627029 |
| Right.I.III | 0.4443561362626351 | 0.0534185978240889 | 0.4303842723632287 | 0.2341473959750984 |
| Left.IV | 0.1387215158826864 | 0.0887677789676057 | 0.1126594290426918 | 0.072184238286071 |
| Right.IV | 0.2444420753731383 | 0.2088656083063521 | 0.3755200403809818 | 0.4476945196633967 |
| Left.V | 0.1309639615103313 | 0.2051495509357963 | 0.1121195547510823 | 0.1519569884860628 |
| Right.V | 0.1749972875997732 | 0.2105343283365087 | 0.2591848172560462 | 0.1173199706651302 |
| Vermis.VI | 0.7989686179506514 | 0.1329076375002257 | 0.1725483987805643 | 0.2830563551980944 |

| | | | | |
|---|---|---|---|---|
| Left.VI | 0.0474471226677328 | 0.0372840416980599 | 0.1431185380052244 | 0.2625980367705439 |
| Right.VI | 0.0474445911384196 | 0.0930594156738161 | 0.1477866402775408 | 0.1268494210775236 |
| Vermis.VII | 0.3179221703634557 | 0.1104327081665734 | 0.3137062704263127 | 0.3837910897636055 |
| Left.Crus.I | 0.2332865978533471 | 0.0275709477084509 | 0.0771298260353733 | 0.0649471373815028 |
| Left.Crus.II | 0.0538921343127936 | 0.1337651941112516 | 0.090628333449256 | 0.0598173323524315 |
| Left.VIIB | 0.0474445890693246 | 0.0343002060255756 | 0.0795250469406859 | 0.3123712283584055 |
| Right.Crus.I | 0.2385420929083691 | 0.0227723345902464 | 0.0654989265866837 | 0.1358079778671702 |
| Right.Crus.II | 0.3200431906873144 | 0.0562952020203151 | 0.1649194671614344 | 0.2692267873915467 |
| Right.VIIB | 0.0763102492812305 | 0.044808006865792 | 0.0533969660540423 | 0.0594018617459417 |
| Vermis.VIII | 0.3848341898786535 | 0.0617976912468601 | 0.0856073872249935 | 0.3816238248461943 |
| Left.VIIIA | 0.0740491000245 | 0.1591943177000737 | 0.4345621147220758 | 0.0996473216397055 |
| Left.VIIIB | 0.0743578331536732 | 0.0796614370022465 | 0.0896224251753463 | 0.139310839095064 |
| Right.VIIIA | 0.0739805904763146 | 0.1117639526412772 | 0.2096108571503737 | 0.4510172384803733 |
| Right.VIIIIA | 0.2241211050665462 | 0.1772422527396371 | 0.1643880446035618 | 0.0858283442097823 |
| Vermis.IX | 0.3715342532129269 | 0.2228288707028127 | 0.4994196013970247 | 0.4342606209882206 |
| Left.IX | 0.3077880624651939 | 0.2384967390597595 | 0.1013737246374847 | 0.1726100994328301 |
| Right.IX | 0.7003252307892722 | 0.1284474606158129 | 0.1453264954067891 | 0.2676656429975352 |
| Vermis.X | 0.3426720617443785 | 0.1343426144215897 | 0.6322928462074843 | 0.7294897428443491 |
| Left.X | 0.2100041950704971 | 0.1677490193427299 | 0.1250586690628136 | 0.1151073384029664 |
| Right.X | 0.2523613692579468 | 0.1703553273462949 | 0.293825160621735 | 0.5436233192615738 |

### c) Feature Importance of AD in task-based atlas

| Features | z_variance_Importance | z_median_Importance | z_positive_Importance | z_negative_Importance |
|---|---|---|---|---|
| 1: Left-hand presses/ motor planning/ interference resolution | 0.1723879842638989 | 0.0378107070958859 | 0.1536545303078618 | 0.186171170181322 |
| 2: Right-hand presses/ motor planning/ divided attention | 0.2127379800928299 | 0.0669235205811555 | 0.0438389459977101 | 0.1683291898064463 |
| 3: Saccades/visual working memory/visual letter recognition | 0.0797685646151132 | 0.082403482903861 | 0.2964273115584614 | 0.0870677606816604 |
| 4: Action Observation/divided attention/motor planning | 0.1325136409034549 | 0.1336039486911354 | 0.0595689555905307 | 0.0830618890910562 |
| 5: Divided attention/active maintenance/mental arithmetic | 0.1321639284062264 | 0.0729348661187938 | 0.0502647782880701 | 0.2277403057400999 |
| 6: Divided attention/verbal fluency/active maintenance | 0.192675279119409 | 0.0584321038223232 | 0.0426868441976423 | 0.070960195589288 |

| | | | | |
|---|---|---|---|---|
| 7: Narrative/ emotion processing/ language processing | 0.1409660992522493 | 0.1280490085317932 | 0.0592766341248923 | 0.2097957376421692 |
| 8: Word comprehension/ language processing/ narrative | 0.1805993512469107 | 0.1081308870008455 | 0.1114493018004904 | 0.1595327995649596 |
| 9: Verbal Fluency/word comprehension/mental arithmetic | 0.1723340430881938 | 0.1039380458403984 | 0.194118351413804 | 0.0580215299391173 |
| 10: Autobiographical recall/visual letter recognition/interference resolution | 0.2255145465920331 | 0.0545689213656628 | 0.0497435109106365 | 0.3818487940709116 |

### d) Feature Importance of AD in resting-state atlas

| Features | z_variance_Importance | z_median_Importance | z_positive_Importance | z_negative_Importance |
|---|---|---|---|---|
| 1: Visual A | 0.2179481630038701 | 0.2643264344910783 | 0.2322423574859014 | 0.1653800852600826 |
| 2: Visual B | 0.1220079531355557 | 0.3913191855938182 | 0.1936276143981396 | 0.2025810170668418 |
| 3: Somatomotor A | 0.2971973562450903 | 0.1112931230836025 | 0.1638353218418841 | 0.3011810903123099 |
| 4: Somatomotor B | 0.3464375257663533 | 0.0369264143930065 | 0.15945489598039 | 0.0680785258219411 |
| 5: Dorsal Attention A | 0.2061189882742481 | 0.2418932238432946 | 0.3460428788652459 | 0.366301461912134 |
| 6: Dorsal Attention B | 0.1678571536047239 | 0.0422837488522962 | 0.1159748930579681 | 0.1981806964448982 |
| 7: Salience/Ventral Attention A | 0.163120556055422 | 0.0619259943332651 | 0.0767087222754238 | 0.0320516400107169 |
| 8: Salience/ Ventral Attention B | 0.3066140927821512 | 0.1459366865756281 | 0.0780117494537807 | 0.0904671012456375 |
| 9: Limbic B | 0.1962744721195644 | 0.1208060245631507 | 0.1618462390828123 | 0.1735794450080115 |
| 10: Limbic A | 0.1273059735445137 | 0.1959001772271265 | 0.277492980359558 | 0.4658049165169229 |
| 11: Control A | 0.4181582178153883 | 0.2576317134218707 | 0.262768600536473 | 0.1980517090372676 |
| 12 Control B | 0.1420711072828552 | 0.1067295793748155 | 0.088786578660715 | 0.397312179620422 |
| 13: Control C | 0.1381991545847651 | 0.127682350700831 | 0.0570385532331342 | 0.2013134743096441 |
| 14: Default A | 0.3353469340192553 | 0.1550704199884994 | 0.7232498805391607 | 0.1666918469666871 |
| 15: Default B | 0.4441656451194087 | 0.1307579044170973 | 0.3478969599053547 | 0.4355760323939139 |
| 16: Default C | 0.1631205557472006 | 0.1457421497603914 | 0.0833773470555149 | 0.2202171297988936 |
| 17: Temporal Parietal | 0.1130536638037718 | 0.204733557688208 | 0.0629177568156981 | 0.116644039763395 |
